# Atlas of Quality of Life in Binocular Visual Field Loss: A Comprehensive Study

**DOI:** 10.64898/2026.06.02.26354170

**Authors:** Luo Song, Lucy Zha, Anagha Lokhande, Julia Seungjoo Baek, Jinghan Wang, Mengyu Wang

**Author notes:** Equal contribution as co-first authors (Luo Song, Lucy Zha, Anagha Lokhande, Julia Seungjoo Baek, Jinghan Wang). Correspondence to: Mengyu Wang, Ph.D. Schepens Eye Research Institute 20 Staniford Street, Boston, MA 02114, USA. Funding Support: NIH R01 EY036222 and NIH P30 EY003790.

## Abstract

**Purpose:** To quantify the binocular integrated visual field (IVF) loss patterns with archetypal (AT) analysis and their associations with patients’ Quality of Life (QoL).

**Design:** Retrospective study.

**Participants:** Over 125,000 patients from three datasets from Massachusetts Eye and Ear and Glaucoma Research Network Consortium.

**Methods:** We used: (1) the Glaucoma Research Network excluding the Massachusetts Eye and Ear subset for the binocular archetypal model training (77, 270 IVFs from 77 270 patients), (2) Massachusetts Eye and Ear dataset for demographic correlation analysis (47,965 IVFs from 47,965 patients), and (3) the MEE Quality of Life Survey dataset for QoL correlation analysis (75 IVFs from 75 patients). The whole study was restricted to the most recent VF measurements from each subject and binocular VFs were constructed by the integrated visual field method, which was taking the higher sensitivity at each test location. We first applied archetypal analysis to cluster 24-2 binocular VFs into archetypal patterns. The total number of patterns was determined by the Bayes factor. Pearson’s correlations analyzed the associations between patients demographic information, binocular VF patterns and QoL scores, and the coefficients were set to 0 if p-values corrected by multiple comparisons ≥ 0.05.

**Main Outcome Measures:** A binocular VF archetypal patterns and its relationships with demographic divergences and QoL.

**Results:** We identified 17 binocular VF loss patterns. Patterns with major vision impairment (AT10, AT12, AT13, AT14, and AT17) were more common in older patients, while Black or African Americans exhibited a broader spectrum of visual loss, notably AT5 and AT12, compared to Asian and White counterparts. 81 MEE patients with QoL survey data was analyzed to investigate the impact of demographic and vision-related variables on QoL. Older age and female gender were significantly associated with lower QoL. Binocular central vision loss (AT 5) and total vision loss (AT 12) had a significantly greater impact on QoL than binocular peripheral vision loss (AT 2, AT 5, AT 16).

**Conclusions:** Individuals with central or total vision loss, as well as certain demographic groups, experience a significantly greater impact on quality of life. The quantifications of binocular VF loss patterns by archetypal analysis may help better understand glaucoma’s impact on patients’ quality of life.

## Introduction

Visual field (VF) loss is a defining feature of glaucoma and other optic neuropathies and serves as a principal marker of disease severity and progression. Standard automated perimetry remains the clinical gold standard for VF assessment, with staging and management decisions traditionally based on monocular indices such as mean deviation and pattern deviation maps.^1^ However, monocular VF testing evaluates each eye independently and does not fully reflect real-world visual function, which depends on the integrated use of both eyes. As a result, monocular VF metrics may underestimate the functional burden experienced by patients in daily life.

Prior work has demonstrated that the location and pattern of VF loss carry important clinical, prognostic, and biological significance beyond global severity measures alone.^2^ Using unsupervised machine learning approaches such as archetypal analysis, reproducible patterns of glaucomatous VF loss have been identified, including central defects, hemifield loss, altitudinal defects, and peripheral loss patterns.^3^ These computationally derived archetypes have been shown to correlate with disease progression, visual field reversal phenomena, and genetic risk profiles, supporting their clinical relevance.^4, 5^ Subsequent studies further extended this framework to characterize central VF loss in advanced disease and to identify patterns associated with rapid progression and increased risk of visual disability.^4, 6^

In parallel, a substantial body of literature has established that VF loss is strongly associated with vision-related quality of life (QoL). Population-based and clinic-based studies consistently demonstrate that VF impairment adversely affects daily functioning, mobility, reading ability, social engagement, emotional well-being, and independence.^7, 8^ Importantly, several investigations have shown that central VF loss disproportionately impacts QoL compared with peripheral loss, even when overall VF severity is similar.^9^ Longitudinal analyses further indicate that the location of progressive VF damage predicts subsequent declines in patient-reported outcomes, emphasizing the importance of spatial VF characteristics in determining functional vision.^10^

Because functional vision is inherently binocular, binocular VF assessment provides a more clinically meaningful representation of visual capability than monocular testing alone. Integrated visual field (IVF) methods, which combine paired monocular VFs by selecting the better sensitivity at each test location, have been proposed to approximate binocular vision and better predict real-world performance.^11^ Prior studies using binocular or binocularly derived VF metrics have demonstrated stronger associations with mobility limitations, fall risk, and quality-of-life measures than monocular indices.^12^ Nevertheless, comprehensive characterization of distinct binocular VF loss patterns and their differential impact on QoL remains limited.^13^

Demographic disparities further shape the relationship between VF loss and patient outcomes. Age, sex, race, and ethnicity influence glaucoma prevalence, severity at presentation, and rates of progression. Large cohort studies have shown that Black patients experience a higher risk of severe VF loss and more aggressive disease phenotypes, while sex-based differences in symptom reporting and functional impairment have also been observed.^14, 15^ However, the extent to which these demographic factors interact with specific binocular VF loss patterns to shape quality of life has not been systematically examined.

This study endeavors to bridge the existing research gap by employing a novel approach to analyze the combined binocular visual field of vision loss patients as shown in **Figure 1**. We first utilized the results from standard 24-2 visual field tests to construct an integrated binocular visual field. Subsequently, we applied archetypal analysis to cluster the diverse presentations of binocular visual field loss into 17 distinct patterns. By correlating these archetypal patterns with demographic information and validated quality of life questionnaires, our research aims to provide a deep and comprehensive understanding of how specific types of visual field defects affect the daily lives of individuals with visual field impairment. This approach promises to offer valuable insights for ophthalmologists and optometrists in counseling patients and developing personalized rehabilitation strategies.

**Figure 1.**
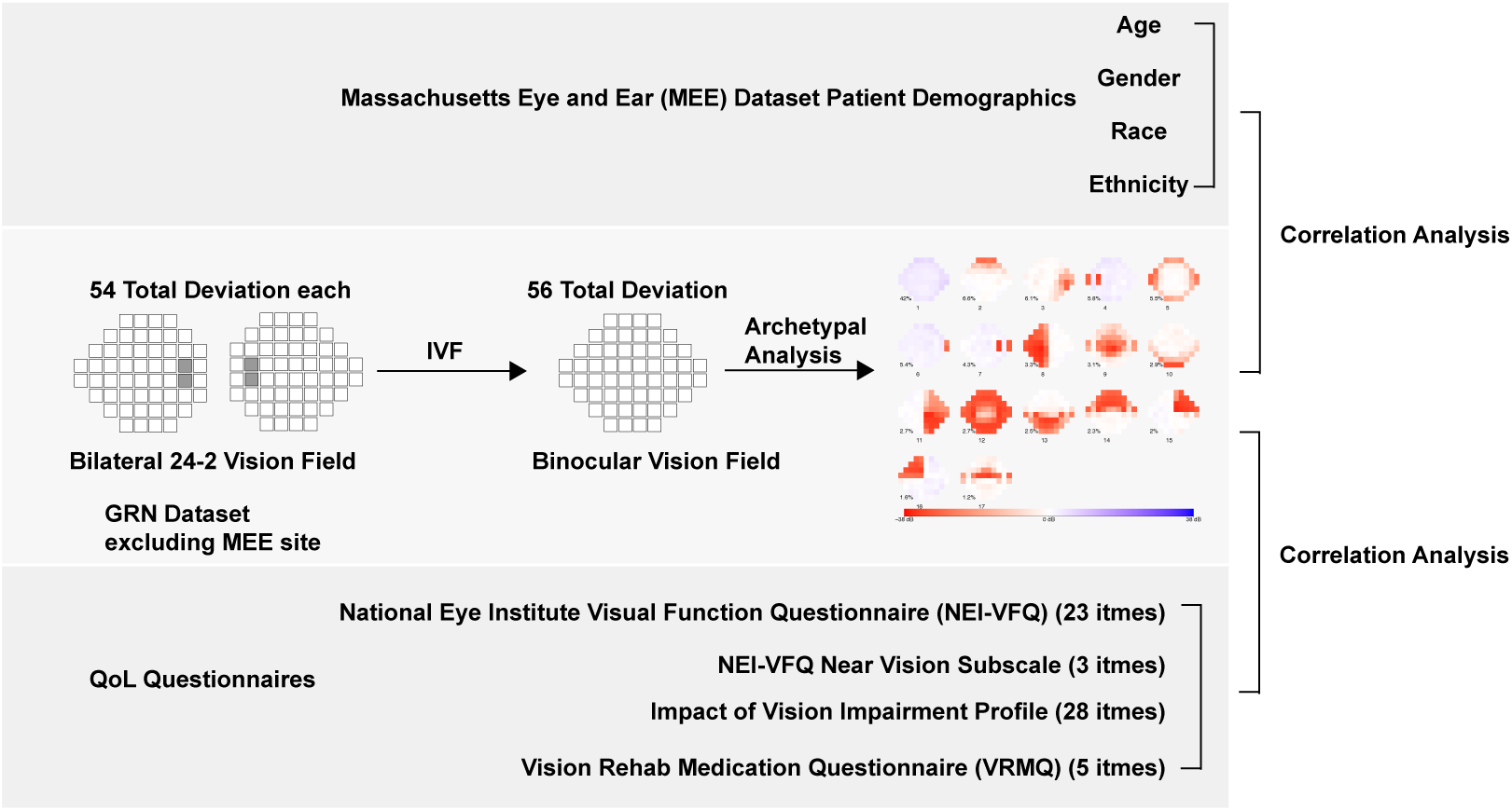
Overview of study: binocular visual field loss and impact on patient quality of life (QoL). This figure illustrates the correlation analyses performed in the study: (A) the relationship between Massachusetts Eye and Ear clinical dataset and archetypal models of visual field loss, and (B) the relationship between QoL questionnaire scores and these archetypal models.

## Methods

### Subjects and Data

Three different datasets were used for our model training and correlation analysis (Figure 1). Only reliable 24-2 VFs (false-negative rates ≤ 20%, false-positive rates ≤ 20%, and fixation losses ≤ 33%) were included in data analyses; otherwise, there were no exclusion criterion for age or refractive error. All binocular vision field in this study was constructed for each patient using the integrated visual field (IVF) method. This was calculated by comparing each patient’s recorded paired monocular VFs and taking the higher sensitivity at each test location, which results in total 56 TD values as a binocular VF.

The first dataset is from the Glaucoma Research Network (GRN) and was used for our binocular archetype model training. We only included the last VF from each pair of eyes measured on the same day for the model training to avoid biasing the training data that have long VF follow-ups. We excluded the VFs from Massachusetts Eye and Ear (MEE), which was the second dataset, used for subsequent correlation analysis for demographic information. Here, we used reliable 24-2 VFs and corresponding demographic information from the MEE dataset. The third dataset, from the three questionnaire surveys done from part of MEE rehab patients. As shown in **Supplemental Table 1**, The questionnaire including 25-item National Eye Institute Visual Function Questionnaire (NEI-VFQ) with additional 3-item Near Vision Subscale, 28-item Impact of Vision Impairment Profile, and 6-items Vision Rehab Medication Questionnaire (VRMQ). For NEI-VFQ, question 15 and 16 was excluded from study due to missing from record. For VRMQ, the first question was ruled out due to limited responses.

### Archetype Analysis

We employed archetype analysis to identify distinct binocular VF loss patterns from GRN dataset. To generate patterns, the total deviation (TD) values of IVFs were used as input data for the archetype analysis, which was performed 20 times, with archetypes having the lowest residual sum of squares (RSS) (Figure 1). RSS values for archetype numbers ranging from 2 to 20 were computed independently. The optimal pattern number was determined using a penalty term of logN (where N is the pattern count) and considered the largest drop of RSS × logN compared to (N-1) archetype patterns, guided by Bayes factor value from the t-test. After obtaining the optimal number of binocular archetypal patterns, each archetypal pattern coefficient was calculated for entire dataset, and patient VFs with top three coefficients of each archetypal were plotted as example accordingly.

### Rasch Analysis for Questionnaires

To provide interval-level measures of QoL, we performed Rasch analysis for individual questionnaires and combined them together. The R package used for analysis was “TAM.”^16^ Due to the polytomous nature of the questionnaire data, in which rating scales differed across items, we applied the Rasch partial credit model (PCM). Questionnaire responses were first recoded because TAM requires the lowest response category to be coded as 0. To ensure consistency, a score of 0 was defined as the worst QoL, with higher scores representing better QoL. For example, a score of 0 for loneliness indicated feeling lonely all the time, whereas a score of 0 for shopping indicated stopping shopping because of eyesight. The theta weighted likelihood estimate (WLE) was used as the QoL score for the corresponding models. Item fit was assessed using Infit Mean Square (MNSQ) statistics, with values close to 1.0 indicating good fit to the Rasch model; values substantially greater than 1.0 suggested underfit (unexpected variability), whereas values substantially less than 1.0 suggested overfit (overly predictable responses).

### Correlation Analysis for Binocular Visual Pattern with Demographics and QoL Questionnaires

Building on our prior work which had established impact of demographics on regional VF loss, we used Pearson’s correlation analysis to investigate the relationships between binocular VF patterns and i) patient demographics (age, gender, race, ethnicity) and ii) scores from three Quality of Life (QoL) questionnaires. To account for multiple comparisons, the p-values of questions under the same sets were adjusted using the False Discovery Rate (FDR) method.

To further investigate the influence of demographic factors, the correlation analyses were stratified by gender (male/female) and race (White/Non-White). A separate analysis was also conducted adjusting for age to control for its potential confounding effect. Stratification by ethnicity was not performed due to an insufficient sample size.

## Result

### Subjects and Data

**Table 1** shows the statistical distributions of age, gender, race, ethnicity, spherical equivalent, cylinder, and 24-2 mean deviation for the GRN, MEE, and MEE-QoL Questionnaire datasets. Compared with the MEE dataset, the GRN dataset has similar patient age (62.7 ± 16.5 vs 57.8 ± 18.3 years) and spherical equivalent (2.1 ± 2.7 vs 2.2 ± 2.6 dioptres) and worse mean deviation (-4.6 ± 5.9 vs -3.9 ± 5.5 dB), while the patients from MEE-QoL Questionnaire has worse mean deviation than previous dataset.

**Table 1.** The statistical distributions of age, spherical equivalent, and 24-2 mean deviation for the three datasets used in the study.

| Dataset | GRN | MEE | MEE-QoL |
| --- | --- | --- | --- |
| No. of IVF | 77270 | 47965 | 75 |
| Binocular Spherical Equivalent (Diopters) | 2.1±2.6 | 2.2±2.6 | 2.0±3.5 |
| Binocular Cylinder | 1.5±1.4 | -1.8±0.9 | -1.9±0.7 |
| Binocular 24-2 Mean Deviation (dB) | -4.4±5.1 | -3.6±4.7 | -12.8±9.9 |
| Age (years) | 62.7±16.5 | 57.3±18.3 | 60.8±16.2 |
| Gender (Female %) | / | 28501 (59.4) | 40 (53.3) |
| Race (%) |  |  |  |
| White | / | 34005 (70.9) | 56 (74.7) |
| Black | / | 4672 (9.7) | 7 (9.3) |
| Asian | / | 3009 (6.3) | 1 (1.3) |
| Other or Unknown | / | 6279 (13.1) | 11 (14.7) |
| Hispanic (%) | / | 3026 (6.3) | 2 (2.7) |

Abbreviations used: No. = Number; VF = visual field; GRN = Glaucoma Research Network; MEE = Mass Eye and Ear; MEE-QoL = Mass Eye and Ear Quality of Life Multi-Questionnaire Dataset.

### Binocular Archetypes

We conducted a comprehensive archetype analysis and identified 17 VF patterns that effectively represented the GRN dataset. We named archetypal patterns in descending order of their average archetypal coefficient weights and depicted in **Figure 2**. Archetype 1 (AT 1) was the most prominent, encompassing 42.0% of average archetypal coefficient weights. Conversely, AT 17 was the least common with 1.2%. **Supplemental Table 2 and 3** details the distribution of data across various archetypes in GRN and MEE datasets. Meanwhile, Supplement Figure 1 shows RSS with LogN penalty plots. Qualitatively, the 17 AT patterns captured a broad spectrum of VF abnormalities, including normal VF, peripheral and equatorial VF loss, ring defects, hemianopia, quadrantanopia, central and paracentral defects, altitudinal loss, and near-total VF loss with central tunnel preservation. AT 1 (normal VF) was the most prevalent pattern (42.0%), whereas AT 17 (superior paracentral VF loss) was the least common (1.2%). Detailed descriptions and distributions of all archetypes are provided in **Supplemental Table 4**.

**Figure 2.**
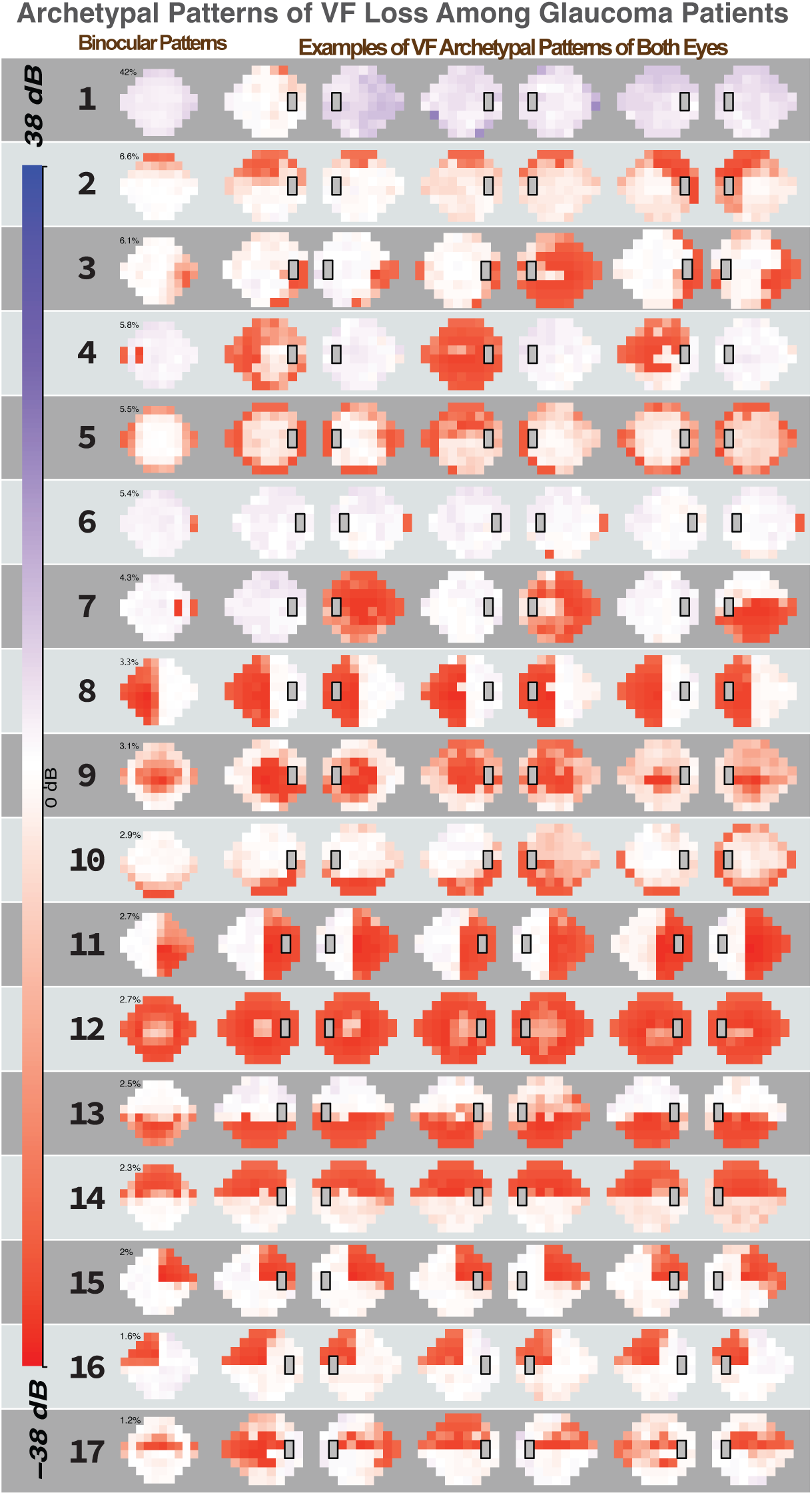
Vision field defect plot of binocular patterns. The binocular archetypal patterns show visual field loss patterns ranging from early to late-stage. The annotation above each plot is the label of plots that represent the membership, while upper left annotation indicates the relative weights. Patient examples of both eyes for each archetypal patterns were given.

### Rasch Analysis for Questionnaires

Traditional statistical methods often assume interval-level data, which poses challenges when analyzing ordinal questionnaire responses. To address this and convert the ordinal scores into interval-level measurements, we applied Rasch analysis. As shown in **Supplemental Table 5**, The analysis evaluated a combined pool of 60 items across 780 participants. Person separation reliability (PSR) was excellent for the combined item set (0.96), the IVI CERA (0.95), and the NEI VFQ 25 (0.94). The Near Vision and Medication Rehab subscales also demonstrated acceptable reliability (PSR = 0.79 for both). Targeting between person ability and item difficulty was optimal for the Near Vision (-0.04), IVI CERA (0.07), and NEI VFQ 25 (0.11) questionnaires. However, the Medication Rehab subscale showed a targeting value of 0.99 (Mean Person Ability SE = 0, Mean Item Difficulty SE = -0.99), indicating that these items were relatively easy for this specific participant sample (N = 208).

Overall, the items demonstrated good fit to the Rasch model. The Infit Mean Square (MNSQ) values for the NEI VFQ 25, Near Vision, and IVI CERA all fell within acceptable thresholds. Only two items exhibited poor fit: one within the overall combined set (Infit MNSQ up to 1.84) and one within the Medication Rehab subscale (Infit MNSQ up to 1.92).

### Correlation Analysis for Demographics

As shown in **Figure 3**, aging is the strongest predisposition to abnormal binocular vision (AT1, -0.24). Meanwhile, it was associated with most binocular VF loss patterns except superior peripheral loss (AT2; R: -0.04), right inferior peripheral loss (AT3, not significant), left and right hemianopia (AT8, AT11, not significant), and left and right superior quadrantanopia (AT15 and 16, not significant). Overall, Hispanic (-0.05), Black (-0.13, -0.11), and male (-0.04) patients were less likely have normal binocular vision. Compared with White patients, Asian patients also tend to have abnormal binocular vision (-0.02). Meanwhile, compared with White and Asian patients, Black patients were more likely to have near total vision loss (AT 12; Rs: 0.1 and 0.08) and peripheral ring loss (AT 5; Rs: 0.07 and 0.08). Black patients were also more likely to have a superior altitudinal loss (AT 14; R: 0.06) than White patients. This result confirmed in **Supplemental Table 3** where there is a higher percentage of African American patients showing peripheral ring loss, inferior altitudinal loss and left superior altitudinal loss (16.5%, 25.3%, 21%). Compared with White patients, Black patients had increased risk for binocular vision loss associated with brain damage, including left nasal step(0.02), right central scotoma (0.01), right superior altitudinal loss (0.01), left superior altitudinal loss (0.06), right outer quadrantanopia (0.02). Apart from age itself, all other mentioned correlations were adjusted with age. Adjusted P values were significant for all the correlations mentioned.

**Figure 3:**
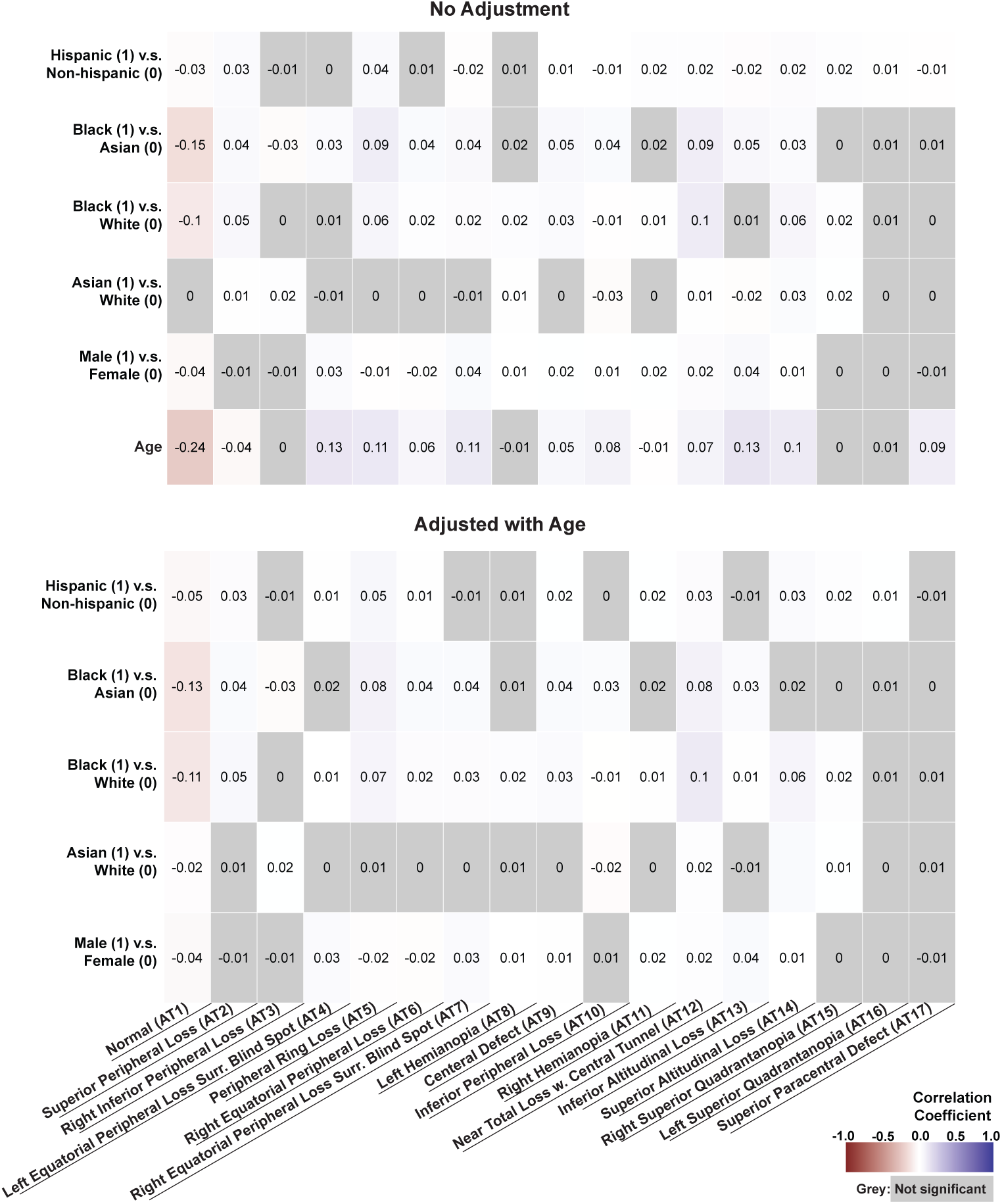
Correlation Between Binocular Visual Field (VF) Patterns and Quality-of-Life Measures Before and After Age Adjustment.

### Correlation between Binocular VF Patterns and Quality of Life

As shown in **Figure 4**, patients with normal binocular vision either have no significant or positive association with high scores. However, binocular central defect (AT 9), near total vision loss with central tunnel (AT 12) and superior altitudinal loss had a significantly greater impact across various QoL Questions.

**Figure 4:**
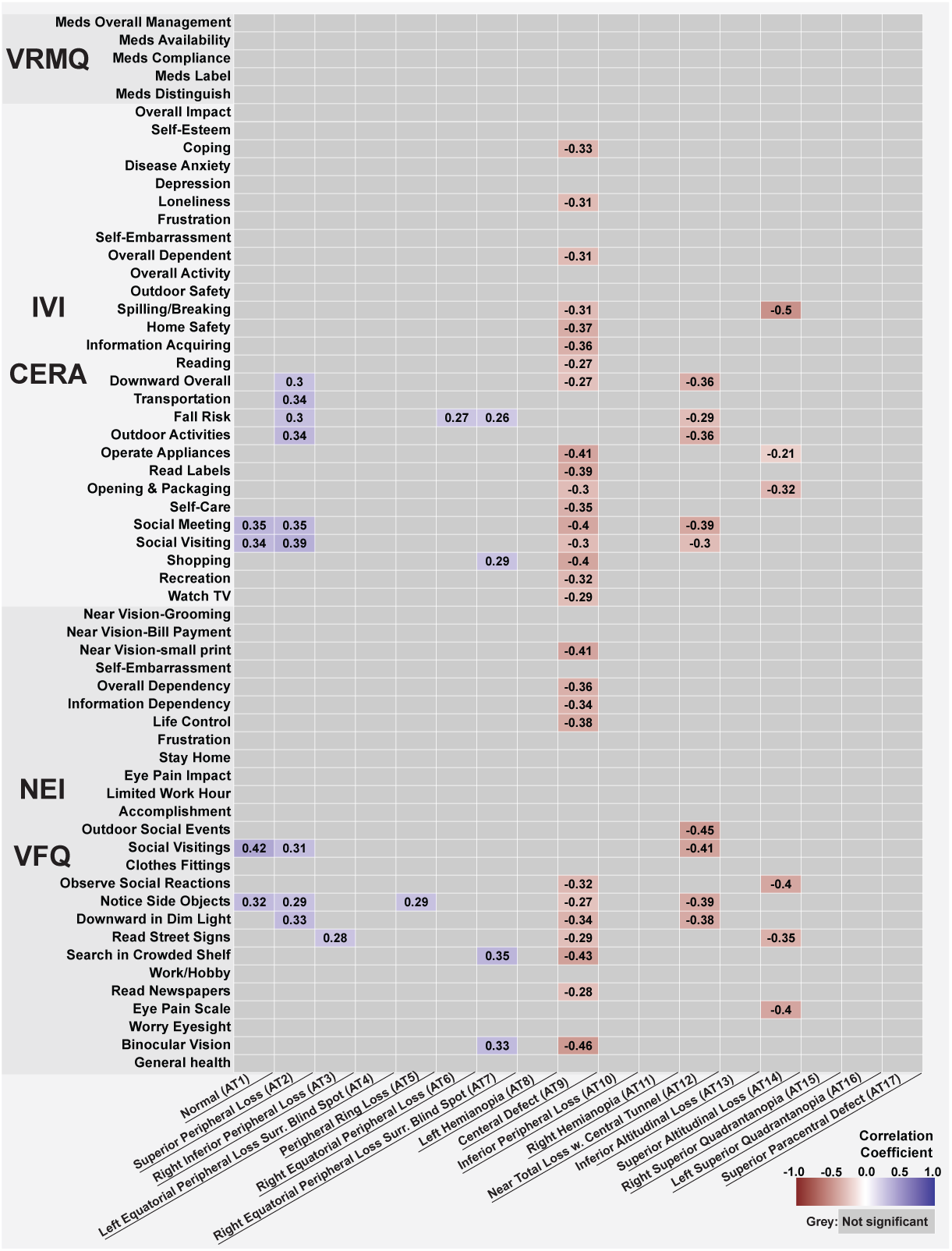
Correlation Between Binocular Visual Field (VF) Patterns and Quality of Life. Heatmap illustrates correlation coefficients between binocular VF archetypal patterns and individual items from vision-related quality-of-life instruments, including the Veterans RAND Medication Questionnaire (VRMQ), Impact of Vision Impairment (IVI), IVI-CERA, and NEI-VFQ-25. Positive correlations (blue) indicate better reported quality of life with the corresponding VF pattern, whereas negative correlations (red) indicate worse quality of life. Numeric values denote correlation coefficients. Grey cells represent non-significant associations.

Central Defect (AT9) demonstrates the most pervasive negative impact, exhibiting widespread significant negative correlations across the IVI, CERA, and NEI VFQ subscales. Specifically, a Central Defect strongly hinders practical and fine-detail tasks, correlating negatively with searching in a crowded shelf (R = -0.43), operating appliances (R = -0.41), and reading small print (R = -0.41). It also impacts psychological well-being, showing negative correlations with life control (R = -0.38) and coping (R = -0.33). Meanwhile, daily activities like watching TV (R = -0.29), Recreation (R = -0.32), shopping (R = -0.4) were impacted. Conversely, patients with Normal VFs (AT1) or Superior Peripheral Loss (AT2) exhibit positive correlations primarily in social and mobility domains, such as social engagements (R = 0.31 to 0.42), social meetings (R = 0.35), and outdoor activities (R = 0.34), indicating preserved function in these areas.

Other severe visual field defects exhibit targeted negative impacts on specific daily activities. Near Total Loss with Central Tunnel (AT12) severely restricts environmental and social interaction, showing strong negative correlations with outdoor social events (R = -0.45), social visiting (R = -0.41), and noticing side objects (R = - 0.39). Similarly, Superior Altitudinal Loss (AT14) is strongly associated with difficulties in physical interaction and observation, correlating negatively with spilling/breaking items (R = -0.50), the eye pain scale (R = -0.40), and observing social reactions (R = -0.40). Interestingly, Left Hemianopia (AT8) presents isolated positive correlations with specific tasks, including searching a crowded shelf (R = 0.35) and binocular vision (R = 0.33).

Other peripheral loss patterns (AT3–AT7) demonstrate only sparse, weak-to-moderate positive correlations with isolated variables such as fall risk (R = 0.26 to 0.27) and noticing side objects (R = 0.29).

### Sex Stratified Correlation Analysis for Binocular VF Patterns

The stratified correlation analysis by sex reveals some differences, as shown in **Figure 5**. For normal vision and mild impaired binocular vision, female patients demonstrate more positive responses, while male patients tends to show no significant correlation. In near total vision loss with central tunnel, female patients tends to have impaired social meetings and visiting (R: -0.58 and -0.43), while male tends to show no significant correlation except impaired opening and packaging (R: -0.53). Conversely, for central defect, female shows no significant correlation, while male shows significant impaired clothes fitting (R: -0.39), read street sign (R: -0.66), searching in crowded shelf (R: -0.48).

**Figure 5:**
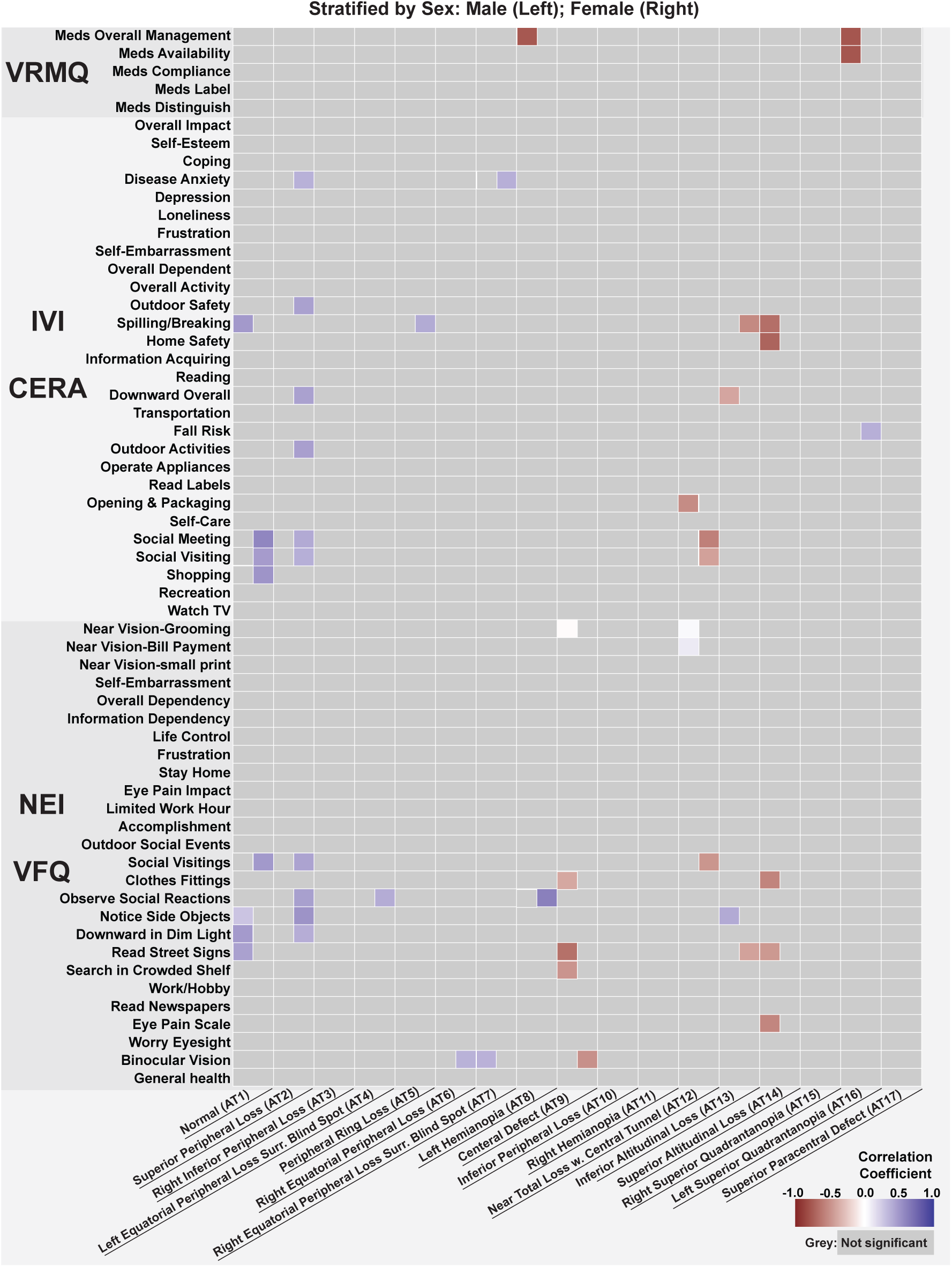
Sex-Stratified Correlation Between Binocular Visual Field (VF) Patterns and Quality-of-Life Measures.

### Race Stratified Correlation Analysis for Binocular VF Patterns

The correlation analysis, stratified by race (White vs. Non-White), reveals distinct demographic patterns in how different visual field defects impact quality-of-life domains, as shown in **Figure 6**. Central Defect (AT9) and Near Total Loss with Central Tunnel (AT12) demonstrate the most pervasive negative impacts, particularly among Non-White patients. For Non-White individuals, a Central Defect (AT9) exhibits broad negative associations across the IVI, CERA, and NEI VFQ subscales, such as Coping (R: -0.69) and Overall Dependency (R: -0.32). Near Total Loss with Central Tunnel (AT12) shows even more severe, targeted negative impacts predominantly for Non-White patients, yielding very strong negative correlations similar as Central Defect. In contrast, White patients exhibit fewer broad negative correlations in these specific defect categories, though they do show strong negative correlations across medication management in the VRMQ (e.g., Meds Compliance and Meds Label, R: -0.89 and - 0.76), as well as isolated moderate negative impacts in AT9 (e.g., Overall Dependency, R: -0.34). Conversely, several VF patterns demonstrate significant positive correlations, highlighting stark racial disparities in functional adaptation or self-reporting. Most notably, Right Superior Quadrantanopia (AT15) presents strong positive correlations exclusively for Non-White patients across multiple functional and social NEI VFQ domains, including Notice Side Objects (R: 0.89), Read Street Signs (R: 0.7), and Observe Social Reactions (R: 0.85).

**Figure 6:**
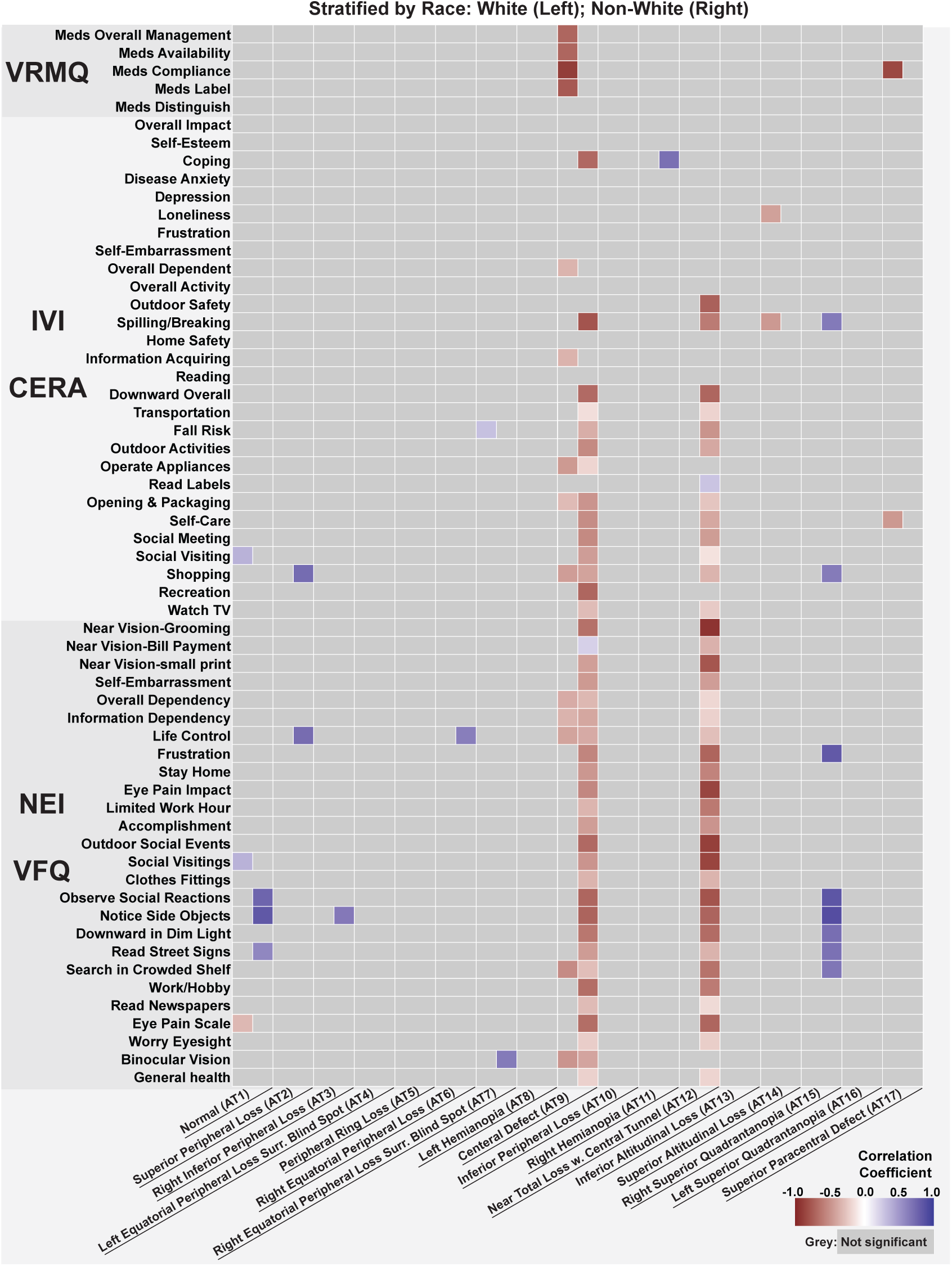
Race-Stratified Correlation Between Binocular Visual Field (VF) Patterns and Quality-of-Life Measures. Heatmaps display correlation coefficients between binocular VF archetypal patterns (columns) and individual items from vision-related quality-of-life instruments, including the Veterans RAND Medication Questionnaire (VRMQ), Impact of Vision Impairment (IVI), IVI-CERA, and NEI-VFQ-25 (rows), stratified by race (White participants shown on the left; non-White participants shown on the right). Color intensity reflects the magnitude and direction of associations (red = negative, blue = positive), with numeric values indicating correlation coefficients. Grey cells denote non-significant associations.

## Discussion

This study provides a comprehensive characterization of binocular VF loss using archetypal analysis applied to integrated visual fields from more than 100,000 patients. We identified 17 distinct binocular VF loss patterns spanning the full spectrum of visual impairment, from normal vision to near-total loss, and showed that these patterns are differentially associated with patient-reported quality of life and demographic factors. Three main findings emerged from this work. First, central binocular defects (AT9) and near-total loss with central tunnel vision (AT12) were associated with the most severe and widespread QoL impairment, affecting functional capacity, social participation, and emotional well-being. Notably, central loss was associated with QoL decrements comparable in magnitude to those observed with near-total vision loss. This finding underscores the functional importance of central binocular vision, even when peripheral fields are relatively preserved.

Second, peripheral VF loss patterns, although they contribute to overall VF severity, were associated with relatively limited QoL impact. This suggests that global severity indices alone may not fully capture the functional burden experienced by patients. Third, we observed significant demographic disparities in both the distribution of binocular loss patterns and their QoL consequences. Black or African American patients showed a broader spectrum of severe binocular loss patterns, and other non-White, non-Black patients experienced more widespread QoL associations for comparable VF deficits.

Sex-stratified analyses further revealed distinct QoL profiles. Female patients reported greater social and emotional impact from near-total loss, whereas male patients showed more task-specific impairment associated with central defects. Together, these findings highlight the importance of considering both demographic context and the spatial pattern of binocular VF loss when assessing functional impact in glaucoma.

From a clinical perspective, these results support several actionable recommendations: prioritizing preservation of central binocular visual field in glaucoma management, incorporating binocular VF spatial pattern assessment alongside traditional monocular indices, using pattern-specific risk profiles to guide early referral to vision rehabilitation services, and adopting equity-informed screening strategies for populations at higher risk of severe binocular loss.

QoL analyses showed that binocular central defects and near-total loss with central tunnel vision were associated with the greatest impairments across functional, social, and emotional domains. Notably, central binocular loss produced QoL decrements comparable to near-total vision loss, underscoring the disproportionate functional impact of central involvement even when peripheral vision remains. These findings align with prior studies demonstrating that VF location predicts QoL beyond global severity.^10^

Distinct binocular patterns exhibited pattern-specific QoL profiles. Superior altitudinal loss was associated with greater emotional distress and psychosocial burden, whereas inferior altitudinal loss was more strongly linked to mobility- and task-related limitations, including fall risk and reading difficulty. These differences highlight that patients with similar overall VF severity may experience markedly different functional challenges depending on the spatial distribution of binocular loss. In contrast to prior work, we uniquely provide VF loss archetype-specific metrics of QoL, which can guide clinicians in individualized counseling and represent an important step toward personalized medicine in glaucoma.^17, 18^

Clinically, these results support prioritizing central binocular function in disease management, using binocular spatial patterning to identify patients at high risk for functional decline, and initiating early referral to vision rehabilitation when high-impact patterns emerge. Pattern-specific insights may further guide tailored counseling and rehabilitation strategies.

Significant demographic disparities were observed. Older age was associated with more severe binocular loss, and female sex was associated with lower QoL, consistent with prior literature. Black or African American patients demonstrated a broader distribution of severe binocular loss patterns, including near-total and peripheral ring loss, aligning with established evidence of higher glaucoma burden in this population. Race-stratified analyses showed that non-White participants experienced broader and more severe QoL impacts for the same VF patterns, likely reflecting the combined effects of disease severity, access to care, and contextual factors.^19, 20^

While most prior studies have evaluated global visual field severity or selected visual field regions,^15, 21, 22^ our study provides the first large-scale, data-driven characterization of binocular visual field loss patterns and their differential associations with quality of life across demographic subgroups. Importantly, our results demonstrate that the functional consequences of binocular visual field loss are not fully explained by overall severity. Patients with central binocular defects experienced QoL impairment comparable to that observed in near-total visual field loss despite substantially greater preservation of peripheral vision. These findings suggest that the spatial configuration of binocular visual field damage may be as important as, or more important than, aggregate measures of visual field loss in determining functional burden. By moving beyond traditional severity metrics and examining the full landscape of binocular visual field patterns, our study provides a more nuanced understanding of how glaucoma affects patients’ daily lives.

Furthermore, whereas previous studies have largely evaluated overall associations between visual field loss and QoL,^22, 23^ our analysis reveals substantial heterogeneity in these relationships across demographic groups. The broader and more severe QoL impacts observed among non-White participants, together with sex-specific differences in functional, social, and emotional outcomes, suggest that the lived experience of glaucoma is shaped not only by the location of visual field loss but also by demographic and social factors. These findings highlight the importance of incorporating equity-focused and patient-centered approaches into glaucoma assessment and management.

This study has limitations. First, the QoL analysis included only 81 patients from a single tertiary referral center, limiting statistical power and generalizability, particularly for race- and sex-stratified analyses. Some large correlations observed in small subgroups should therefore be interpreted cautiously. Second, the cross-sectional design precludes causal inference, and longitudinal studies are needed to determine whether changes in binocular VF patterns predict subsequent QoL decline. Third, QoL was assessed using self-reported questionnaires, which may be influenced by cultural and linguistic differences. Fourth, integrated visual fields approximate binocular function but do not fully capture binocular summation or suppression. Finally, analyses were adjusted only for age and may be subject to residual confounding.

Future studies should validate binocular archetypal patterns in larger, prospectively enrolled cohorts with longitudinal QoL follow-up. Further work should also evaluate whether incorporating binocular VF pattern information into clinical decision-support tools improves patient outcomes and quality of care.

## Conclusion

Our study characterizes binocular VF loss patterns using archetypal analysis and demonstrates their differential associations with vision-related quality of life (QoL) and demographic factors. We demonstrate an important use case for artificial intelligence in quantifying QoL disparities in glaucoma, extending the bounds of its utility in glaucoma beyond the conventional applications in image processing and diagnostics.^24^ By integrating large-scale clinical VF data with patient-reported outcomes, we extend prior monocular VF pattern analyses and provide a more functionally relevant, personalized medicine framework for understanding how spatial patterns of binocular vision loss translate into real-world disability.

Heatmaps show correlation coefficients between binocular visual field (BVF) archetypes and individual items from vision-related quality-of-life questionnaires. The top panel presents unadjusted correlations, while the bottom panel shows correlations after adjustment for age. Cell values indicate correlation coefficients, with color intensity reflecting the magnitude and direction of association (red = negative correlation, blue = positive correlation). Greyed cells indicate non-significant associations.

## Data Availability

All data produced in the present study are available upon reasonable request to the authors

