## Supplemental Tables & Figures for "Atlas of Quality of Life in Binocular Visual Field Loss: A Comprehensive Study"

Supplemental Table 1. Questionnaire details

| Questionnaire | Question ID | Short Description | Content |
| --- | --- | --- | --- |
| National Eye Institute Visual Function Questionnaire  (NEI-VFQ)  Scale: 0 - 4 | Q1 | General Health | In general, would you say your overall health is? |
|  | Q2 | Subjective Binocular Vision | At the present time, would you say your eyesight using both eyes (with glasses or contact lenses, if you wear them) is excellent, good, fair, poor, or very poor or are you completely blind? |
|  | Q3 | Worry Eyesight | How much of the time do you worry about your eyesight? |
|  | Q4 | Eye Pain | How much pain or discomfort have you had in and around your eyes (for example, burning, itching, or aching)? |
|  | Q5 | Read Newspapers | How much difficulty do you have reading ordinary print in newspapers? |
|  | Q6 | Work/Hobby | How much difficulty do you have doing work or hobbies that require you to see well up close, such as cooking, sewing, fixing things around the house, or using hand tools? |
|  | Q7 | Search in Crowded | Because of your eyesight, how much difficulty do you have finding something on a crowded shelf? |
|  | Q8 | Read Street Signs | How much difficulty do you have reading street signs or the names of stores? |
|  | Q9 | Downward in Dim | Because of your eyesight, how much difficulty do you have going down steps, stairs, or curbs in dim light or at night? |
|  | Q10 | Notice Side Objects | Because of your eyesight, how much difficulty do you have noticing objects off to the side while you are walking along? |
|  | Q11 | Observe Social Reactions | Because of your eyesight, how much difficulty do you have seeing how people react to things you say? |
|  | Q12 | Clothes Fittings | Because of your eyesight, how much difficulty do you have picking out and matching your own clothes? |
|  | Q13 | Social Visiting | Because of your eyesight, how much difficulty do you have visiting with people in their homes, at parties, or in restaurants? |
|  | Q14 | Outdoor Social Events | Because of your eyesight, how much difficulty do you have going out to see movies, plays, or sports events? |
|  | Q17 | Accomplishment | Do you accomplish less than you would like because of your vision? |
|  | Q18 | Limited Work Hour | Are you limited in how long you can work or do other activities because of your vision? |
|  | Q19 | Eye Pain Impact | How much does pain or discomfort in or around your eyes, for example, burning, itching, or aching, keep you from doing what you'd like to be doing? |
|  | Q20 | Stay Home | I stay home most of the time because of my eyesight, |
|  | Q21 | Frustration | I feel frustrated a lot of the time because of my eyesight. |
|  | Q22 | Life Control | I have much less control over what I do, because of my eyesight. |
|  | Q23 | Information Dependency | Because of my eyesight, I have to rely too much on what other people tell me. |
|  | Q24 | Overall Dependency | I need a lot of help from others because of my eyesight |
|  | Q25 | Self-embarrassment | I worry about doing things that will embarrass myself or others, because of my eyesight. |
|  | N1 | Near Vision – small print | Wearing glasses, how much difficulty do you have for reading the small print in a telephone book, on a medicine bottle, or on legal forms? Would you say: |
|  | N2 | Near Vision Subscale – Bill Payment | Because of your eyesight, how much difficulty do you have figuring out whether bills you receive are accurate? |
|  | N3 | Near Vision Subscale - Grooming | Because of your eyesight, how much difficulty do you have doing things like shaving, styling your hair, or putting on makeup? |
| Impact of Vision Impairment Profile  Scale: 0 - 3 | I1 | Watch TV | Your ability to see and enjoy T.V? |
|  | I2 | Recreation | Taking part in recreation activities such as bowling, walking or golf? |
|  | I3 | Shopping | Shopping? (finding what you want and paying for it) |
|  | I4 | Social Visiting | Visiting friends or family? |
|  | I5 | Social Meeting | Recognizing or meeting people? |
|  | I6 | Self-care | Generally looking after your appearance? (face, hair, clothing etc.) |
|  | I7 | Opening & Packaging | Opening packaging? (for example, around food, medicines) |
|  | I8 | Read Labels | Reading labels or instructions on medicines? |
|  | I9 | Operate Appliances | Operating household appliances and the telephone? |
|  | I10 | Outdoor Activities | How much has your eyesight interfered with getting about outdoors? (on the pavement or crossing the street) |
|  | I11 | Fall Risk | In the past month, how often has your eyesight made you go carefully to avoid falling or tripping? |
|  | I12 | Transportation | In general, how much has your eyesight interfered with travelling or using transport? (bus & train) |
|  | I13 | Downward Overall | Going down steps, stairs, or curbs? |
|  | I14 | Reading | Reading ordinary size print? (for example, newspapers) |
|  | I15 | Information Acquiring | Getting information that you need? |
|  | I16 | Home Safety | Your general safety at home? |
|  | I17 | Spilling/Breaking | Spilling or breaking things? |
|  | I18 | Outdoor Safety | Your general safety when out of your home |
|  | I19 | Overall Activity | In the past month, how often has your eyesight stopped you doing the things you want to do? |
|  | I20 | Overall Dependent | In the past month, how often have you needed help from other people because of your eyesight? |
|  | I21 | Self-Embarrassment | Have you felt embarrassed because of your eyesight? |
|  | I22 | Frustration | Have you felt frustrated or annoyed because of your eyesight? |
|  | I23 | Loneliness | Have you felt lonely or isolated because of your eyesight? |
|  | I24 | Depression | Have you felt sad or low because of your eyesight? |
|  | I25 | Disease Anxiety | In the past month, how often have you worried about your eyesight getting worse? |
|  | I26 | Coping | In the past month how often has your eyesight made you concerned or worried about coping with everyday life? |
|  | I27 | Self-esteem | Have you felt like a nuisance or a burden because of your eyesight? |
|  | I28 | Overall Impact | In the past month, how much has your eyesight interfered with your life in general? |
| Vision Rehab Medication Questionnaire (VRMQ)  Scale: 0 - 3 | M1 | Medication Help | Does anyone help you take or manage your medications? |
|  | M2 | Meds Distinguish | Using your current glasses, how much difficulty telling one medication from another? |
|  | M3 | Meds Label | Using your current glasses, how much difficulty reading your medication labels? |
|  | M4 | Meds Compliance | Using your current glasses, how much difficulty taking your medications? |
|  | M5 | Meds Availability | Using your current glasses, how much difficulty buying or ordering your medications from the pharmacy? |
|  | M6 | Meds Overall Management | Using your current glasses, how much difficulty managing your medication routine in general? |

Supplemental Table 2. Distribution of data across 17 archetypes for GRN dataset

| Archetype | Binocular 24-2 Mean Deviation (dB) | Binocular Spherical Equivalent (Dioptres) | Binocular Cylinder | Age (years) |
| --- | --- | --- | --- | --- |
| 1 (normal VF) | -2 ± 2.2 | 2.2 ± 2.6 | 1.5 ± 1.4 | 60.3 ± 16.5 |
| 2 (superior peripheral) | -7.9 ± 3.3 | 2.6 ± 2.5 | 1.6 ± 1.3 | 68.9 ± 15.1 |
| 3 (right inferior peripheral) | -10.7 ± 3.8 | 1.5 ± 3.6 | 1.5 ± 1.5 | 71.1 ± 14.6 |
| 4 (left equatorial peripheral) | -9.2 ± 3.6 | 2 ± 2.6 | 1.6 ± 1.5 | 68 ± 15 |
| 5 (peripheral ring) | -10.6 ± 3.9 | 2.7 ± 2.5 | 1.6 ± 1.4 | 68.9 ± 15.6 |
| 6 (right equatorial peripheral) | -4.7 ± 2.5 | 2.5 ± 2.6 | 1.5 ± 1.4 | 66.2 ± 15.4 |
| 7 (right equatorial peripheral VF loss surrounding bind spot) | -9.9 ± 3.4 | 2 ± 2.5 | 1.6 ± 1.4 | 68.9 ± 14.2 |
| 8 (left hemianopia) | -15.5 ± 4.1 | 2 ± 3.1 | 1.4 ± 1.5 | 65.8 ± 18.3 |
| 9 (central defect) | -16.8 ± 4.9 | 1.5 ± 3 | 1.5 ± 1.6 | 71.3 ± 15.4 |
| 10 (inferior peripheral) | -11.2 ± 3.9 | 2.1 ± 2.4 | 1.7 ± 1.3 | 68.9 ± 15.9 |
| 11 (right hemianopia) | -15.5 ± 3.9 | 1.9 ± 2.8 | 2 ± 1.2 | 67.2 ± 16.3 |
| 12 (near total loss with central tunnel) | -21.7 ± 3.8 | 1.8 ± 2.5 | 1.9 ± 1.5 | 67.5 ± 17.1 |
| 13 (inferior altitudinal VF loss) | -17.1 ± 4.1 | 1.8 ± 2.5 | 1.7 ± 1.4 | 71.8 ± 14.1 |
| 14 (superior altitudinal VF loss) | -16.4 ± 4 | 1.8 ± 2.7 | 1.7 ± 1.3 | 71 ± 13.3 |
| 15 (right superior quadrantanopia) | -13.6 ± 4 | 1.9 ± 2.8 | 1.8 ± 1.7 | 70.3 ± 14.8 |
| 16 (left superior quadrantanopia) | -12.3 ± 4 | 1.9 ± 2.4 | 1.6 ± 1.5 | 71 ± 14.4 |
| 17 (superior paracentral) | -13.1 ± 3.9 | 1.9 ± 2.6 | 1.6 ± 1.2 | 72.8 ± 12.1 |

Supplemental Table 3. Distribution of data across 17 archetypes for MEE dataset

| Archetype | Binocular 24-2 Mean Deviation (dB) | Binocular Spherical Equivalent (Dioptres) | Binocular Cylinder | Age (years) | Female (%) | White (%) | Black (%) | Asian (%) | Hispanic (%) |
| --- | --- | --- | --- | --- | --- | --- | --- | --- | --- |
| 1 (normal VF) | -1.9 ± 2.1 | 2.1 ± 2.6 | -1.8 ± 0.9 | 55.8 ± 18.1 | 21477  (60.1) | 25539(71.5) | 3262(9.1) | 2324(6.5) | 2227(6.2) |
| 2 (superior peripheral) | -8 ± 3.3 | 2.9 ± 2.1 | -1.8 ± 0.8 | 65.7 ± 15.9 | 410(57.6) | 429(60.3) | 124(17.4) | 55(7.7) | 59(8.3) |
| 3 (right inferior peripheral) | -10.2 ± 3.9 | 2.1 ± 3.2 | -2 ± 1 | 65 ± 17.1 | 144(56.9) | 179(70.8) | 33(13) | 11(4.3) | 24(9.5) |
| 4 (left equatorial peripheral) | -9.2 ± 3.8 | 2.5 ± 2.6 | -1.9 ± 0.9 | 65.4 ± 16.1 | 1094  (52.6) | 1446(69.5) | 224(10.8) | 123(5.9) | 132(6.3) |
| 5 (peripheral ring) | -11.3 ± 4 | 3.1 ± 2.3 | -1.6 ± 0.8 | 64.6 ± 17.2 | 309(63.8) | 304(62.8) | 80(16.5) | 29(6) | 53(11) |
| 6 (right equatorial peripheral) | -4.3 ± 2.6 | 2.8 ± 2.4 | -1.9 ± 1 | 64.1 ± 16.7 | 1121  (61.7) | 1236(68) | 222(12.2) | 99(5.4) | 137(7.5) |
| 7 (right equatorial peripheral VF loss surr. bind spot) | -10.1 ± 3.6 | 2.4 ± 2.4 | -2 ± 1 | 65.5 ± 16.7 | 883(51.8) | 1171(68.8) | 203(11.9) | 105(6.2) | 78(4.6) |
| 8 (left hemianopia) | -15.2 ± 3.4 | 2.5 ± 2.8 | -1.8 ± 1.2 | 60.8 ± 17.8 | 137(50) | 205(74.8) | 27(9.9) | 13(4.7) | 11(4) |
| 9 (central defect) | -16.3 ± 5.1 | 2.2 ± 3.7 | -2.3 ± 1.1 | 68.1 ± 18.3 | 81(43.3) | 133(71.1) | 21(11.2) | 12(6.4) | 7(3.7) |
| 10 (inferior peripheral) | -10.6 ± 4 | 2.6 ± 2.2 | -1.7 ± 0.6 | 62.6 ± 18.1 | 125(51.2) | 174(71.3) | 26(10.7) | 15(6.1) | 11(4.5) |
| 11 (right hemianopia) | -15.1 ± 4.1 | 2.4 ± 2.6 | -1.4 ± 1 | 61.6 ± 17.8 | 98(41) | 180(75.3) | 19(7.9) | 7(2.9) | 16(6.7) |
| 12 (near total loss with central tunnel) | -24.3 ± 4.7 | 2.6 ± 3.1 | -2 ± 1 | 65.1 ± 17.7 | 285(54.2) | 263(50) | 133(25.3) | 23(4.4) | 56(10.6) |
| 13 (inferior altitudinal VF loss) | -16.6 ± 4.1 | 2.7 ± 2.3 | -1.8 ± 1 | 69.3 ± 14.9 | 132(38.2) | 251(72.5) | 45(13) | 11(3.2) | 8(2.3) |
| 14 (superior altitudinal VF loss) | -16.8 ± 4.2 | 2.6 ± 2.8 | -2 ± 0.9 | 71.3 ± 13.4 | 138(48.3) | 149(52.1) | 60(21) | 32(11.2) | 24(8.4) |
| 15 (right superior quadrantanopia) | -11.4 ± 3.7 | 2.7 ± 3 | -1.7 ± 0.7 | 60.7 ± 19.7 | 57(50.9) | 72(64.3) | 14(12.5) | 6(5.4) | 8(7.1) |
| 16 (left superior quadrantanopia) | -10.8 ± 3.8 | 2.8 ± 1.9 | -1.5 ± 0.8 | 65.4 ± 19.1 | 60(51.7) | 86(74.1) | 12(10.3) | 5(4.3) | 7(6) |
| 17 (superior paracentral) | -13 ± 4.1 | 2.6 ± 2.4 | -2.2 ± 1 | 72.3 ± 12.1 | 128(66.3) | 139(72) | 20(10.4) | 15(7.8) | 7(3.6) |

Supplemental Table 4. Clinical Characteristics and Relative Prevalence of the 17 Binocular Visual Field Archetypes Identified in the GRN Dataset

| **AT Pattern** | **Description** | **Percent of Coefficient Weights** |
| --- | --- | --- |
| 1 | Normal VF | 42.00% |
| 2 | Superior peripheral VF loss | 6.60% |
| 3 | Right inferior peripheral VF loss | 6.10% |
| 4 | Left equatorial peripheral VF loss surrounding blind spot | 5.80% |
| 5 | Peripheral ring VF loss | 5.50% |
| 6 | Right equatorial peripheral VF loss | 5.40% |
| 7 | Right equatorial peripheral VF loss surrounding blind spot | 4.30% |
| 8 | Left hemianopia | 3.30% |
| 9 | Central defect | 3.10% |
| 10 | Inferior peripheral VF loss | 2.90% |
| 11 | Right hemianopia | 2.70% |
| 12 | Near total loss with central tunnel | 2.70% |
| 13 | Inferior altitudinal VF loss | 2.50% |
| 14 | Superior altitudinal VF loss | 2.30% |
| 15 | Right superior quadrantanopia | 2.00% |
| 16 | Left superior quadrantanopia | 1.60% |
| 17 | superior paracentral VF loss | 1.20% |

Supplement Table 5. Rasch analysis results of QoL across different Questionnaire

| Questionnaire | N items | N persons | Mean Person Ability SE | Mean Item Difficulty SE | Targeting | Infit MNSQ range | items with poor fit | Person Separation Reliability |
| --- | --- | --- | --- | --- | --- | --- | --- | --- |
| All | 60 | 780 | -0.16 (0.03) | -0.35 (0.08) | 0.19 | 0.8 - 1.84 | 1 | 0.96 |
| NEI VFQ 25 | 23 | 755 | 0 (0.03) | -0.11 (0.13) | 0.11 | 0.81 - 1.32 | 0 | 0.94 |
| Near Vision | 3 | 624 | 0 (0.05) | 0.04 (0.44) | -0.04 | 0.91 - 1.1 | 0 | 0.79 |
| IVI CERA | 28 | 756 | 0.03 (0.04) | -0.04 (0.11) | 0.07 | 0.84 - 1.34 | 0 | 0.95 |
| Medication Rehab | 6 | 208 | 0 (0.06) | -0.99 (0.24) | 0.99 | 0.72 - 1.92 | 1 | 0.79 |

Supplemental Figure 1.

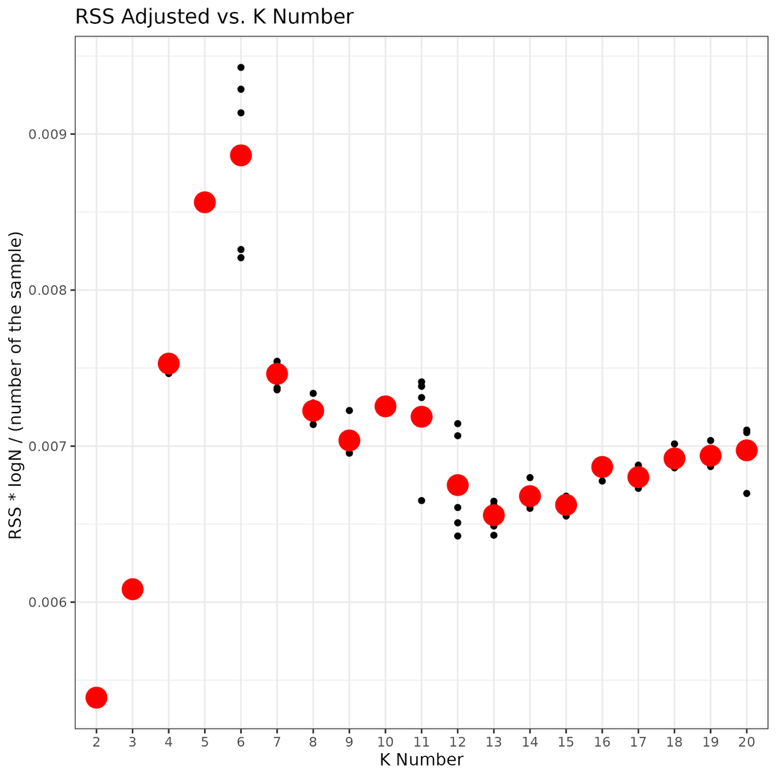
